# Rest-activity and circadian rhythm parameters relate to cognition and disability outcomes in multiple sclerosis

**DOI:** 10.64898/2026.08.31.26361636

**Authors:** Katrin Wolfova, Charles White, Nasim Montazeri Ghahjaverestan, Tenzing Choeying, Rodolfo Arevalo, Kaho Onomichi, Levi Davis, Victoria M. Leavitt, Korhan Buyukturkoglu, Claire Riley, Andrew Lim, Philip De Jager

**Author notes:** **Correspondence:** Philip L. De Jager, MD PhD, Weil-Granat Professor of Neurology, Chief, Division of Neuroimmunology, Director, Center for Translational & Computational Neuroimmunology, Director, Multiple Sclerosis Center, Department of Neurology, Deputy Director, Taub Institute for Research on Alzheimer’s Disease and the Aging Brain, Columbia University Irving Medical Center, 630 West 168th Street, P&S12-461, New York, NY10032 USA.

## Abstract

**OBJECTIVE:** To prioritize novel measures of disease progression, we examined whether actigraphy-derived rest-activity rhythm (RAR) parameters relate to cognitive performance as well as disability in a multiple sclerosis (MS) cohort enrolled in a prospective brain donation program.

**METHODS:** RAR parameters were assessed using a wrist actigraphy device (AX3 Axivity Actiwatch, Axivity Ltd.) over two weeks. The primary outcome measure was the Symbol Digit Modalities Test (SDMT, N=222). Secondary outcomes included Brixton Spatial Anticipation Test and self-reported disability. In 76 participants, volumetric measures were derived from repurposed clinical magnetic resonance imaging (MRI) data. We applied linear and logistic regression models, adjusting for age, sex, education, time since MS diagnosis, and body mass index.

**RESULTS:** After correction for multiple comparisons, higher intradaily variability (IV) of RAR and lower relative amplitude were associated with worse SDMT performance; higher IV was also associated with greater odds of disability. A broader set of RAR parameters was associated with disability measure. No MRI parameters were related to RAR in the subset of individuals with available MRI data, although we note suggestive associations with hippocampal and choroid plexus volumes warranting further investigation.

**INTERPRETATION:** More robust circadian rhythms were related to better cognition. These results highlight the utility of actigraphy and its more nuanced measures beyond the simple summaries of activity levels that quantitate the extent of motor disability. Selected RAR features may be an effective non-invasive approach to capture clinically relevant quantitative measures of brain function for persons with MS.

## INTRODUCTION

Cognitive impairment is estimated to affect 45-70% of people with multiple sclerosis (MS), most commonly assessed with the Symbol Digit Modalities Test (SDMT).^1^ Greater physical activity has been associated with better cognitive performance in MS,^2^ yet the total activity volume alone offers only partial insight into daily movement patterns. Actigraphy-derived rest-activity rhythm (RAR) parameters describe how activity is distributed over the 24-hour cycle, capturing when activity occurs, how robustly it separates from rest, and how consistently that pattern repeats across days. These metrics are particularly relevant in clinical populations such as MS, for whom impaired mobility or fatigue may limit the ability to sustain prolonged activity; in addition, sleep fragmentation impairs a key natural restorative process, further altering activity-rest patterns that can become disrupted in neurodegenerative diseases such as MS.

Emerging evidence shows that people with MS exhibit increased activity fragmentation and reduced variability in active bout length compared to healthy controls,^3^ although the contribution of motor impairment is unclear. Further, accelerometry has detected differences between MS types: compared with relapsing-remitting MS, progressive MS is associated with lower activity, greater within-day fragmentation, and less day-to-day stability of rest-activity patterns.^4^ Whether RAR relates to cognition in MS has not been examined, however, studies in community-dwelling older adults demonstrated that greater fragmentation of both rest and activity are linked to lower levels of cognitive performance, specifically in the perceptual speed, semantic memory, working memory, and visuospatial abilities domains.^5^ Alterations in circadian rhythm amplitude (the day-night difference in activity) have also been linked to aging and are associated with faster cognitive decline and a higher dementia risk.^6–8^

Abnormal rest-activity rhythm has also been related to structural brain differences in regions relevant for cognition. In cognitively unimpaired adults aged ≥ 50, weaker and more fragmented RAR were associated with smaller medial temporal region volumes,^9^ and in older adults with objective and/or subjective cognitive impairment, greater fragmentation was associated with reduced cortical thickness.^10^ In MS, accelerometry-derived activity fragmentation has recently been shown to predict subsequent reduction in deep gray matter and thalamic volumes and disability progression.^11^ The extent to which rest-activity rhythm disruptions relate to cognitive performance, disability, and brain structure in people with MS remains insufficiently understood.

Here, we introduce a new **Resource** for the MS community: the MS Snapshot cohort. It is a longitudinal study of persons with MS that includes prospective brain donation. Participants are recruited nationwide in the United States and characterized with longitudinal cognitive and actigraphy evaluations. All data are available to external investigators through an application process. The cross-sectional analyses presented here extend earlier work and examine whether actigraphy-derived rest-activity rhythm parameters are associated with cognitive performance in MS. The primary cognitive outcome was SDMT. Secondarily, we evaluated whether rest-activity rhythm parameters associated with SDMT performance were also related to executive functioning, measured by total errors on the Brixton Spatial Anticipation Test, and to disability measured by the Patient-Determined Disease Steps (PDDS). These analyses will inform longitudinal studies evaluating whether actigraphy can predict cognitive and motor decline in MS.

## METHODS

### Study design and setting

The MS Snapshot Study is an ongoing prospective cohort study of adults with MS or other neuroinflammatory diagnosis designed to characterize clinical, behavioural, and biological factors associated with disease progression. Participants are recruited either in person at the Columbia MS Center (New York, NY, USA) or remotely across the United States. Recruitment began in 2019 and is ongoing. Participants enrolled remotely are part of the associated Brain Bank program and provide consent for postmortem brain donation for research purposes. Participants recruited through the Columbia MS Center may also opt into the Brain Bank program; however, brain donation is not required for participation. Participants completed a baseline remote visit with a study coordinator, during which a neuropsychological battery was administered. Participants also completed questionnaires assessing demographic characteristics, health history, and patient-reported outcomes, including measures of disability and fatigue (Figure 1). Magnetic resonance imaging (MRI) data from routine clinical care obtained closest to the time of the remote visit were analyzed. As part of the study protocol, participants are asked to wear a wrist-based actigraphy device during the baseline assessment to measure physical activity, circadian rhythm patterns. Exceptions are made when device use is not feasible (e.g., due to allergies, skin sensitivities, or other participant-specific circumstances).

**Figure 1.**
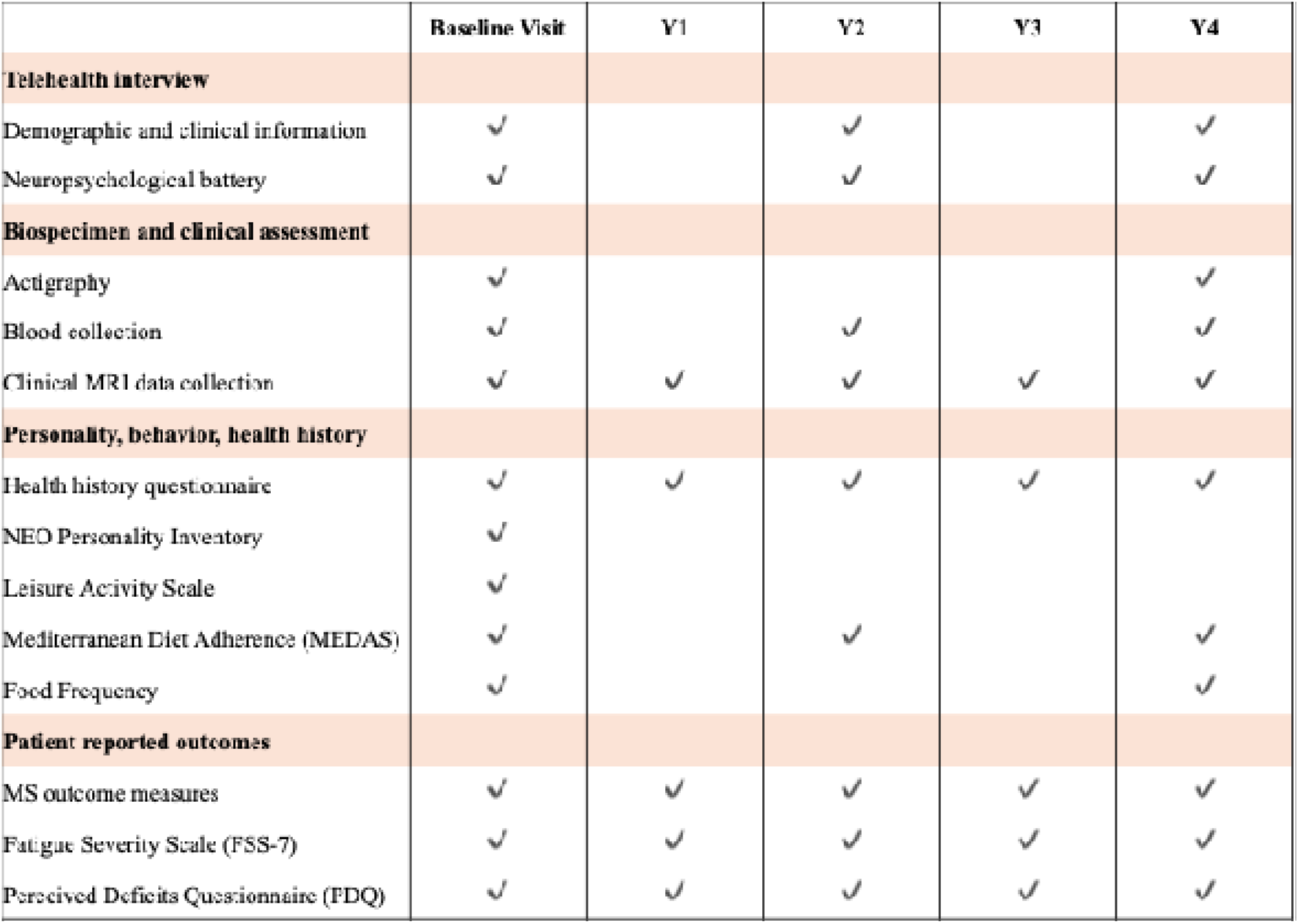
MS Snapshot study schedule

All participants signed informed consent. The study was approved by the Columbia University Medical Center Institutional Review Board.

### Study population

A total of 496 participants completed at least one Snapshot visit by July 2025. Participants with complete data on SDMT, rest-activity rhythm parameters, age, sex, education, time since MS diagnosis and body mass index were included in the primary analytic sample (n=222, Supplementary Figure 1).

In secondary analysis, we further restricted the sample to include those with complete data on Brixton Spatial Anticipation Test (n=197), PDDS (n=208) and on MRI measures (n=76).

### Rest-activity rhythm parameters

Circadian rest–activity rhythms were assessed using wrist actigraphy device. The participants wore an AX3 Axivity Actiwatch (Axivity Ltd.) on their non-dominant wrist for 7 continuous days to measure circadian parameters. Wear time was annotated by visual inspection. Only recordings with at least 7 consecutive days of wear were considered. For recordings with more than 7 consecutive days of wear, we only considered the first 7 full days of recording.

From the three axes of raw accelerometery, a widely used metric, the Euclidean Norm Minus One (ENMO), was first derived.^12^ ENMO is calculated by subtracting 1 g from the Euclidean norm of acceleration and truncating negative values to zero:^12^

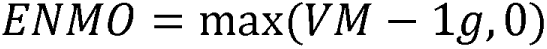

This procedure effectively removes the static gravitational component and yields a gravity-corrected measure of movement intensity expressed in milligravity units (mg).^12^ Lower ENMO values generally reflect sedentary behaviour or rest, while higher values correspond to periods of increased activity.^13^ Additionally, the z-angle (also referred to as wrist angle) was calculated using trigonometric functions based on the relationship between the acceleration axes and the gravitational vector where the z-axis corresponds to the axis positioned perpendicular to the skin surface.^14^ The z-angle has been widely used for the heuristic sleep-detection algorithm developed by van Hees and colleagues without requiring sleep diaries.^14^ From ENMO and z-angle, a sets of indices were extracted.

To model the cyclic 24-hour activity, Cosinor-derived metrics, including Midline Estimating Statistic of Rhythm (mesor), amplitude, acrophase, and Coefficient of Determination (R²), were extracted.^15, 16^ These metrics are derived by fitting a single-component 24-hour cosinor model:

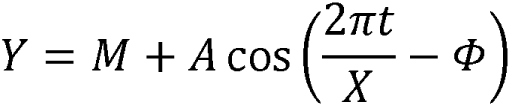

to each cycle of the actigraphy data. In this equation, *Y* represents activity level at time *t*. *M*, *A*, *X* and *ϕ* are mesor, amplitude, the cycle length (24 hours), and the acrophase representing timing of peak activity, respectively. Furthermore, R² was reported as the coefficient of determination of the fitted cosinor model representing the proportion of total variance in the observed times series explained by the model.^17^ Higher R**²** values indicate a stronger, more regular circadian rhythm.^17^ The parameters estimated from this model are described below.^17,18^

Interdaily stability (IS) provides an estimation of how well the 24-hour rhythm is aligned from one day to the next and measures the consistency of activity of patterns across days.^18^ Higher IS values indicate greater day-to-day regularity and a healthier circadian rhythm.^18^ Intradaily variability (IV) provides an estimation of fragmentation within the 24-hour rest-activity rhythm.^18^ Least active 5-hour period (L5) represents the mean activity level during the least active continuous 5-hour period within a 24-hour day and serves as a proxy for nocturnal rest or sleep-related inactivity. Most active 10-hour period (M10) represents the mean activity level during the most active continuous 10-hour period of the day and reflects daytime activity intensity. Relative Amplitude (RA) is calculated using the following equation:

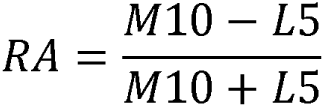

kRA is the average probability of transitioning from a sustained rest state to an activity state. Higher kRA values indicate more frequent rest-to-activity transitions and therefore less consolidated rest. To compute kRA, ENMO values were first aggregated across 15-second epochs and scaled to reproduce activity counts similar to the Actical device (Phillips Respironics).^19^ Sleep time (ST) is the estimated total sleep time per day, calculated based on motionless epochs from wrist angle (z-angle).^14, 20^ Variables with non-normal distributions were rank-normalized, and the remaining variables were standardized using z-scores.

### Clinical outcomes

Clinical outcomes included cognition measured by two tests, the SDMT and the Brixton Spatial Anticipation Test, and disability as defined as score ≥3 using the Patient Determined Disease Scale (PDDS). The PDDS is a validated patient-reported outcome of disability in MS, and correlates strongly with EDSS.^21^

To assess the cognitive dimension, we prioritized the SDMT, which is the most common cognitive outcome used in MS research and clinical trials, and serves as a proxy of current cognitive status.^22^ The Brixton Spatial Anticipation Test is an untimed test that involves detecting a pattern in visuospatial stimuli.^23^ Patterns are governed by logical, mathematical, or directional rules. For SDMT, total correct was the outcome variable; for Brixton, the outcome variable was total number of errors.

### MRI data collection and analysis

MRI scans were acquired within one year of the actigraphy assessment (range: 1–364 days; mean: 117 days) on GE Discovery (3T, n = 48; 1.5T, n = 3) or Siemens (3T, n = 17; 1.5T, n = 8) scanners. All T1-weighted images underwent N4 bias field correction.^24^ White matter lesions were automatically segmented from T1-weighted and FLAIR images using SAMSEG, a module within FreeSurfer,^25^ visually inspected and manually corrected when necessary, and subsequently lesion-filled using FSL.^26^

To account for scanner-related differences, ComBat harmonization (NeuroCombat) was performed using MRI vendor and field strength as batch variables, and harmonized data were used in sensitivity analyses.

### MRI outcomes

Total brain volume, grey matter volume, white matter volume, nucleus accumbens, amygdala, caudate nucleus, hippocampus, thalamus and choroid plexus (CP) volumes were measured using FreeSurfer (version 7.4.1). Volumetric MRI measures were adjusted for intracranial volume (ICV) using a residualization approach.^27^ Specifically, each volume was regressed on ICV, and the resulting residuals were retained as ICV-adjusted volumetric measures for subsequent analyses. For CP volumetry, initial masks were generated using FreeSurfer, followed by an automated correction algorithm and manual editing of all CP masks.^28^

Cortical thickness measures derived from FreeSurfer were combined across individual cortical regions to calculate mean cortical thickness for the major cortical lobes (frontal, parietal, temporal, occipital) and the insular cortex.

### Covariates

Covariates included age at visit (years), sex (male or female), educational attainment (less than high school, high school, some college/associate degree, college degree, or graduate degree and above), time since MS diagnosis (years), and body mass index (BMI; kg/m²). When BMI data were unavailable at a given study visit, the value from the closest available visit was used.

### Analytic strategy

To evaluate the association between rest-activity rhythm parameters and clinical and MRI outcomes, we fitted a series of linear regression models for each rest-activity rhythm parameter and each outcome separately. Models with total errors on the Brixton Spatial Anticipation Test and PDDS were fitted only for rest-activity rhythm parameters that demonstrated significant associations with SDMT performance. Fully adjusted models with SDMT, total errors on the Brixton Spatial Anticipation Test and PDDS as the outcomes were controlled for age, sex, education, time since MS diagnosis, and BMI. Fully adjusted models with brain volumes as the outcomes were controlled for age, sex, time since MS diagnosis and BMI. To account for multiple comparisons across rest-activity rhythm variables, p-values were adjusted using the false discovery rate (FDR) correction. Statistical significance was determined based on FDR-adjusted p-values, with q < 0.05 considered significant.

As a sensitivity analysis, models evaluating associations between rest-activity rhythms and clinical outcomes were additionally adjusted for the mean Fatigue Severity Scale (FSS-7) score to assess whether the observed associations were robust to accounting for fatigue, given its potential relationship with rest-activity patterns, cognition, and disability. As an additional sensitivity analysis, all models with MRI outcomes were repeated using ComBat-harmonized MRI measures to assess the robustness of the results to scanner-related variability.

## RESULTS

The primary analytic sample consisted of 222 participants in the MS Snapshot, a longitudinal cohort of people with MS enrolled in a prospective brain donation program; they have a mean age of 52.3 years (± standard deviation [SD] 13.3) and a predominance of women (78.8%). Most participants were white (82.9%) and college-educated, with 36.5% holding a college degree and 38.7% having completed graduate-level education or above. Just over half resided in the Northeast of the United States (50.9%). The mean disease duration was 15.2 years (± SD 12.2), and 20.7% reported a family history of MS. Neurologic comorbidities were present in 12.6% of participants, and the mean BMI was 27.5 (± SD 6.82). 31.1% had ever smoked, and 59.9% reported alcohol use. The subset of participants included in the MRI outcomes analyses (n=76) was similar in age but included a lower proportion of women (69.7%) and had a shorter mean disease duration (11.2 years, ± SD 9.06). This subset had a higher proportion of participants with graduate-level education (47.4%). Because MRI data were available primarily for participants recruited at the Columbia MS Center, this subset was more heavily concentrated in the Northeast. Participant characteristics are presented in Table 1. All data are available to be repurposed on request.

**Table 1.**
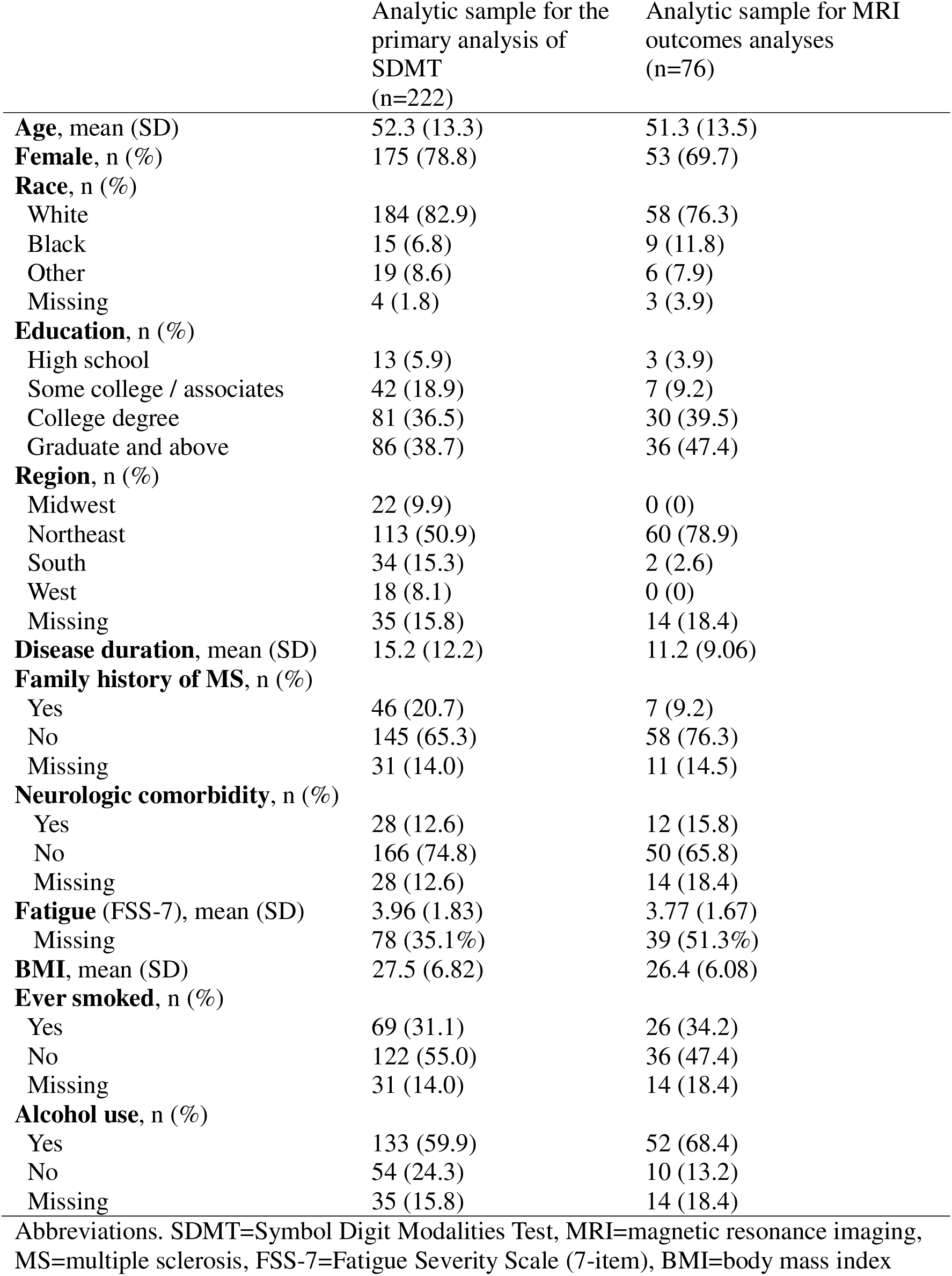
Sample characteristics.

After correction for multiple comparisons using the FDR procedure, several associations between RAR parameters and SDMT performance remained statistically significant. Higher intradaily variability (IV), a measure of heterogeneity in daily activity patterns that reflects poor circadian health, was associated with worse SDMT performance (B = -3.030, 95% CI - 4.590 to -1.470, Table 2, and it was also associated with increased odds of disability derived from the PDDS (OR = 2.036, 95% CI 1.322 to 3.136, Table 2).

**Table 2.**
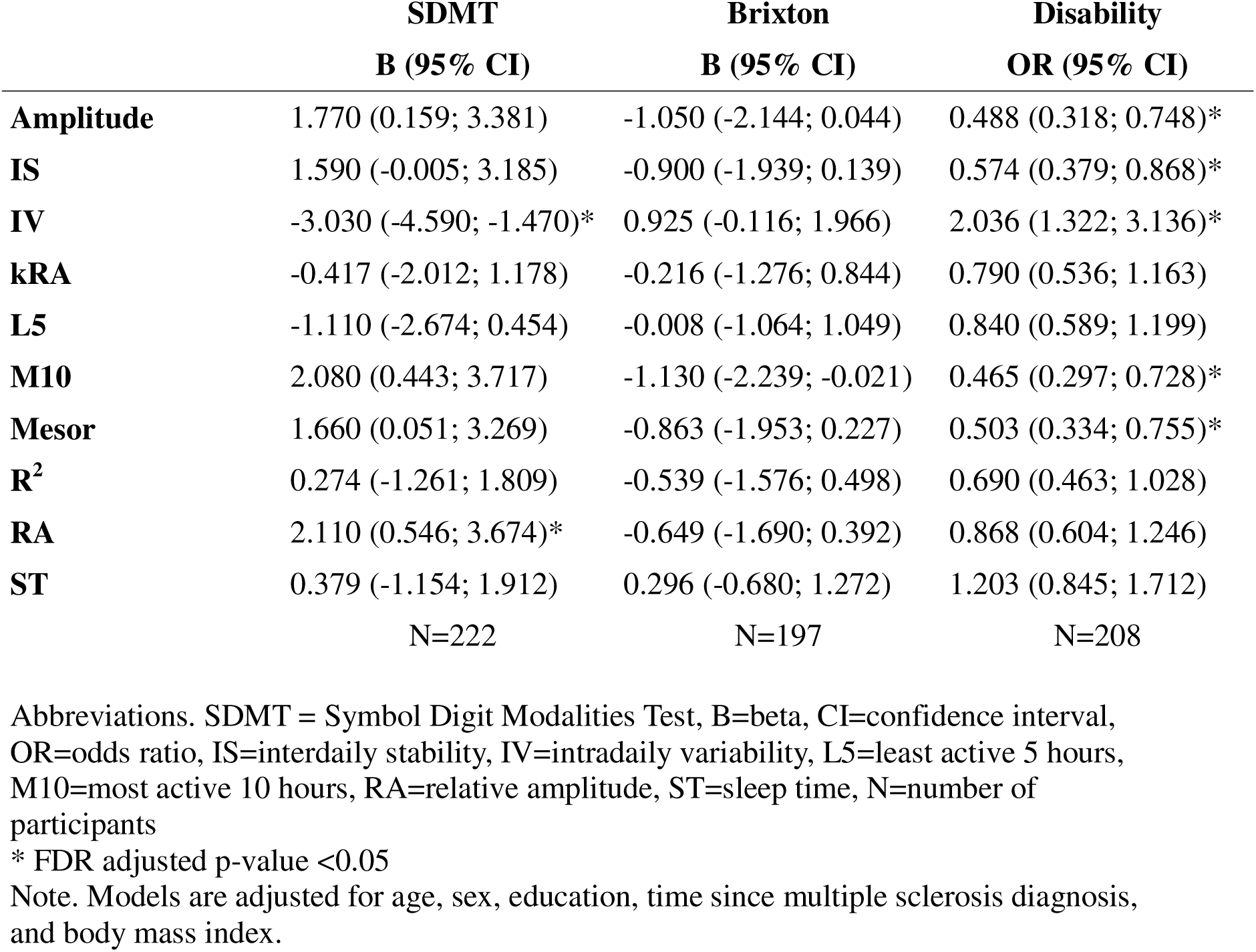
Relationship between RAR parameters and clinical outcomes.

Another RAR parameter that was FDR significant for the SDMT was relative amplitude (RA), a non-parametric metric that captures the relative difference between the most active period and the least active period of activity. In our participants, greater RA was associated with better SDMT performance (B = 2.110, 95% CI 0.546 to 3.674, Table 2) where a higher RA arises from high daily activity and very restful periods of sleep.

In analyses of the PDDS measure, several other parameters were associated with lower odds of disability: (1) higher amplitude of activity (OR = 0.488, 95% CI 0.318 to 0.748) which is related to a more robust circadian rhythm, (2) higher interdaily stability (IS) (OR = 0.574, 95% CI 0.379 to 0.868) which measures the stability of RAR patterns over subsequent days, (3) M10 (OR = 0.465, 95% CI 0.297 to 0.728) the average activity over the most active 10 hours of a daily period, and (4) midline estimating statistic of rhythm (mesor) (OR = 0.503, 95% CI 0.334 to 0.755) which captures a participant activity baseline (Table 2). Of these 4 parameters significantly linked to better PDDS, higher amplitude, M10, and mesor also displayed suggestive evidence (with uncorrected p<0.05 but FDR>0.05) of association with better SDMT performance (Table 2). No correlations were shown to performance on the Brixton. Adjustment for fatigue did not substantially alter the observed associations between rest-activity rhythms and clinical outcomes.

We then explored the relation of our prioritized RAR parameters with MRI-derived volumetric parameters; neither IV nor RA were associated, although IV trended in the expected direction with smaller thalami and other volumes in the context of greater IV. Secondarily, we assessed the relation of all RAR measures to the MRI-derived measures (Figure 2), although the subset of 76 participants with MRI measures is moderate in size. None of these results met an FDR corrected threshold of significance, but several suggestive (uncorrected p<0.05) associations were observed (Figure 2). Higher mesor (B = 351, 95% CI 114 to 588), amplitude (B = 357, 95% CI 118 to 596), and M10 (B = 285, 95% CI 79.2 to 491) were each associated with greater hippocampal volume (Supplementary Table 1).

**Figure 2.**
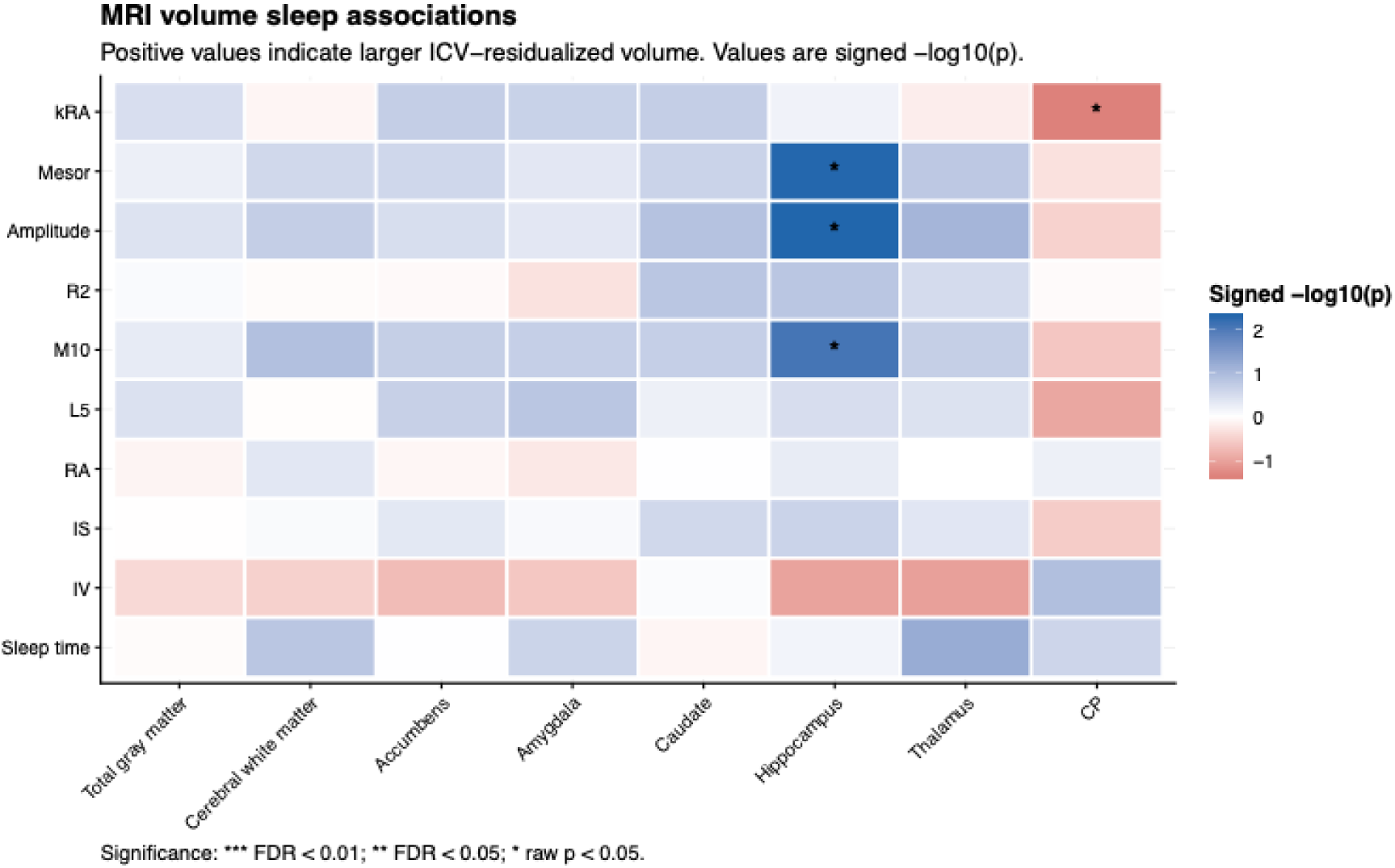
Relationship between RAR parameters and regional brain volumes

Finally, we note a suggestive association of higher kRA, the weighted average probability of transitioning from a state of sustained rest to activity, with smaller choroid plexus volume (B = −144, 95% CI −281 to −7, Supplementary Table 1). We did not find any associations between RAR parameters and cortical thickness.

## DISCUSSION

In this cross-sectional analysis of a prospective study of persons with MS, we found that a more fragmented rest-activity pattern, indexed by higher IV, was associated with worse performance on the SDMT. Conversely, a strong circadian day-night rhythm, reflected by higher RA, was associated with better SDMT performance. Thus, we present evidence that RAR monitoring captures early cognitive changes in MS and could contribute to prioritizing persons for more detailed neuropsychologic testing. This is helpful in the context of individuals with limited access to tertiary, specialized medical care or in widely distributed longitudinal studies such as ours where data are collected remotely. More broadly, we validate a prior study by showing that a more fragmented rest-activity pattern was associated with greater odds of disability measured by the PDDS. Several additional measures of rhythm strength – amplitude, IS, M10, and mesor – were each associated with lower odds of disability, and, in uncorrected analyses that yield suggestive results, with better SDMT performance as well. These results support the use of actigraphy measures to capture quantitative outcomes of CNS function beyond motor measures in MS studies.

The association between RAR fragmentation, worse separation of active periods from rest periods and SDMT performance suggests that circadian integrity may be a more sensitive marker of cognitive dysfunction in MS than just overall activity level. Notably, associations were specific to SDMT and were not observed for executive functioning as measured by the Brixton Spatial Anticipation Test. This diverges somewhat from patterns reported in general aging populations, where RAR disruption has been linked to a broader set of domains, including memory, working memory, and visuospatial ability alongside processing speed.^5^ The relative specificity of our findings to SDMT may reflect the fact that this measure sensitively detects function across a number of cognitive domains including working memory, processing speed, and language functions (as well as non-cognitive domains, such as visual scanning and motor speech function),^29^ making it more sensitive to detect an association with rhythm integrity in a sample of this size. Our persons with MS are also a much younger participant population (mean of 52.3 years of age), which may lead to distinct results from those seen in older individuals and suggest that our results are not simply reflecting age-related changes.

In contrast to our cognitive findings, which were specific to IV and RA, disability showed a broader and more consistent pattern of association across a larger set of RAR parameter we examined. Lower fragmentation of the rest-activity rhythm (IV), higher average activity (mesor), greater daytime activity (M10), day-to-day consistency (IS), and stronger day-night distinction (amplitude) were all associated with lower disability odds. Part of this association likely reflects the direct physical constraints of MS-related disability, which may affect both the ability to engage in sustained activity (and hence lower IV) or affect the ability to engage in regularly scheduled activity (and hance decrease IS). PDDS is a measure of mobility impairment and reduced ambulatory capacity; it would be expected to lower overall activity levels, reduce peak daytime activity, and flatten the contrast between active and rest periods. However, disability was also associated with IV and IS, suggesting that mobility limitations alone do not fully account for these findings. Fatigue, common in MS, may represent additional, partially independent contributors to rhythm disruption in this population.^30^ The persistence of the association between RAR and disability following adjustment for fatigue supports an association that extends beyond the effects of fatigue. Our findings also align with prospective evidence from the Home-based Evaluation of Actigraphy to predict Longitudinal Function in MS study, in which greater activity fragmentation was associated with higher risk of subsequent EDSS-Plus progression,^11^ indicating that fragmentation may not simply be a downstream consequence of impaired mobility. Our study is cross-sectional and cannot comment on causality, but it prioritizes features that we will be evaluating longitudinally in this cohort study.

After correction for multiple comparisons, no associations between RAR parameters and MRI outcomes reached statistical significance. Given that the MRI subsample (n=76) was substantially smaller than our full analytic sample, this null result may reflect limited statistical power rather than a true absence of association and motivates investigation of larger cohorts. As noted above, we did find suggestive evidence that higher mesor, amplitude, and M10 were each associated with greater hippocampal volume. These associations, stemming from age-adjusted analyses, suggest that there may be selective vulnerability of the hippocampus to circadian disruption in persons with MS, which is distinct from the associations of IV and RA with worse SDMT performance. Circadian rhythm dysregulation has been linked to hippocampal vulnerability, and associated deficits in learning and memory.^31^ Our findings are in line with prior work in community-dwelling older adults enrolled in the Baltimore Longitudinal Study of Aging, where lower activity fragmentation was associated with higher temporal lobe white matter volume, a region anatomically adjacent to and structurally connected with the hippocampus. While the observed associations in our data did not survive correction, they raise the possibility of a relationship between rhythm robustness and hippocampal integrity in MS that may be more readily detected in future studies by incorporating hippocampal-dependent measures of episodic memory, in addition to the SDMT or Brixton performance test.

We also found an intriguing, suggestive association of lower kRA, indicating a lower probability of transitions between rest and activity, with greater choroid plexus volume in uncorrected analyses. The choroid plexus has a role in regulation of circadian cerebrospinal fluid dynamics and glymphatic clearance.^32^ Choroid plexus volume has been reported to be enlarged in MS relative to healthy individuals, particularly in relapsing-remitting MS, where it scales with disease duration, lesion burden, and periventricular tissue damage,^33^ and in other neurodegenerative disease like Alzheimer’s disease;^34^ it is thought to be a manifestation of a compartmentalized immune response. This result needs further investigation.

Our study has several strengths. First, we used objective, actigraphy-derived RAR parameters rather than relying on self-reported physical activity and sleep diaries, avoiding recall and social desirability biases common to questionnaire-based measures. Second, actigraphy data were collected over a two-week period, capturing both weekday and weekend activity patterns and providing a more representative picture than shorter monitoring windows. Third, participants in the primary sample were recruited from across multiple US regions, yielding a more geographically and demographically diverse sample than is typical of single-center studies. Finally, we examined a comprehensive set of RAR parameters rather than restricting our analysis to total activity volume, allowing us to characterize more nuanced aspects of daily activity-rest patterns that simple activity counts alone cannot capture. For example, we found different patterns of association with circadian measures than those that emerge with more general measures that capture impaired motor function.

Our study has several limitations. First, the MRI subsample was substantially smaller than our full analytic sample, and the resulting limited statistical power likely explains why associations of RAR parameters with brain volumes did not survive correction for multiple comparisons. Larger studies are needed to confirm these preliminary finding. Based on our uncorrected results, future hypothesis-driven work could focus specifically on hippocampal and choroid plexus volumes as regions of interest. Given the cross-sectional design of this study, we cannot establish the temporal direction of these associations. Rhythm disruption may contribute to worse SDMT performance, for example, through pathways involving fragmented or reduced restorative sleep. Alternatively, worse cognition itself may contribute to a more fragmented activity pattern, as difficulty with working memory and overall cognitive efficiency could manifest as reduced rhythm integrity. Prior longitudinal evidence on this question is limited. In a large study of cognitively intact older adults, daily activity patterns predicted subsequent cognitive decline, but baseline cognition did not predict subsequent changes in activity patterns, arguing against a straightforward reverse-causation explanation in that population.^35^ Whether this pattern holds in MS is unclear.

In conclusion, we introduce a new prospective cohort study of persons with MS that has the rare feature of including brain donation prospectively. Longitudinal RAR parameters are being captured to provide a collection of unbiased ante-mortem measures of CNS function in participants being monitored across the United States. These data – and all other cognitive and MRI data – are available to be repurposed. Our cross-sectional analyses represent a first evaluation of our study strategy and demonstrates that certain RAR features were meaningfully associated with an MS-relevant cognitive measure as well as more general measures of disability. Our secondary analyses highlight several suggestive results that will guide the design of future studies as our sample size increases, particularly in terms of individuals with longitudinal measures. Notable preliminary results link rhythm robustness to hippocampal and choroid plexus volumes. Clinically, accelerometry-derived RAR data could provide physicians with objective information to help identify patients at higher risk of disability and cognitive impairment. These parameters may also help guide personalized exercise recommendations based on a patient’s individual rest-activity pattern. Given that wrist actigraphy is inexpensive and well-tolerated over the course of two weeks of monitoring, it offers a promising tool for longitudinal monitoring of functional status in clinical practice as well as MS research.

## ACKNOWLEDGEMENTS

This study was supported by the National Multiple Sclerosis Society (Project Title: National Multiple Sclerosis Tissue Repository; Award Number: SI-19-1903-33766).

## AUTHOR CONTRIBUTIONS

Conception and design of the study: KW, CW, VL, CL, PLD

Acquisition and analysis of data: CW, KW, TC, RA, KO, AL, NMG, LD, KB

Drafting a significant portion of the manuscript or figures: KW, NMG, KB

## DATA AVAILABILITY

The data generated and analyzed during the current study are not publicly available due to privacy and ethical restrictions. Data may be made available from the corresponding author upon reasonable request and subject to institutional review and approval, completion of the applicable data use agreement (DUA), and any other requirements governing access to the data.

