## Supplement for "Rest-activity and circadian rhythm parameters relate to cognition and disability outcomes in multiple sclerosis"

**SUPPLEMENTARY TABLES**

**Supplementary Table 1**. Relationship between RAR parameters and regional brain volumes

|  | **Gray matter** | **Cerebral white matter** | **Nucleus accumbens** | **Amygdala** | **Caudate nucleus** | **Hippocampus** | **Thalamus** | **Choroid plexus** |
| --- | --- | --- | --- | --- | --- | --- | --- | --- |
|  | **B (95% CI)** | **B (95% CI)** | **B (95% CI)** | **B (95% CI)** | **B (95% CI)** | **B (95% CI)** | **B (95% CI)** | **B (95% CI)** |
| **kRA** | 4,390 (-4,650; 13,400) | -1,710 (-13,300; 9,870) | 32.9 (-16.9; 82.7) | 56.9 (-34.4; 148) | 164 (-84.9; 413) | 52.2 (-177; 282) | -109 (-575; 357) | -144 (-281; -7) |
| **Mesor** | 2,740 (-7,180; 12,700) | 6,960 (-5,580; 19,500) | 31 (-23.5; 85.5) | 37.9 (-62.6; 138) | 168 (-106; 442) | 351 (114; 588) | 361 (-143; 865) | -57.1 (-211; 96.8) |
| **Amplitude** | 4,320 (-5,700; 14,300) | 8,410 (-4,250; 21,100) | 27.1 (-28.2; 82.4) | 38.2 (-63.5; 140) | 214 (-60.4; 488) | 357 (118; 596) | 450 (-55.7; 956) | -78.6 (-234; 76.6) |
| **R2** | 1,070 (-10,300; 12,500) | -932 (-15,500; 13,600) | -5.55 (-68.7; 57.6) | -41.5 (-157; 73.9) | 232 (-79.6; 544) | 209 (-75.2; 493) | 306 (-276; 888) | -12.8 (-190; 165) |
| **M10** | 2,810 (-5,740; 11,400) | 8,570 (-2,170; 19,300) | 30.8 (-16.2; 77.8) | 56.1 (-29.9; 142) | 154 (-81.2; 389) | 285 (79.2; 491) | 285 (-150; 720) | -79.3 (-211; 52.6) |
| **L5** | 4,810 (-5,680; 15,300) | -495 (-13,900; 13,000) | 36.9 (-20.9; 94.7) | 77.6 (-27.8; 183) | 77.2 (-215; 369) | 132 (-133; 397) | 241 (-298; 780) | -133 (-294; 27.7) |
| **RA** | -1,530 (-11,500; 8,450) | 4,830 (-7,830; 17,500) | -7.39 (-62.7; 47.9) | -29.5 (-131; 71.6) | 4.21 (-274; 283) | 79.3 (-172; 330) | 3.21 (-510; 517) | 40.9 (-114; 196) |
| **IS** | -197 (-9,590; 9,190) | 1,200 (-10,800; 13,200) | 19.2 (-32.5; 70.9) | 11.3 (-84; 107) | 144 (-115; 403) | 143 (-92.2; 378) | 188 (-292; 668) | -80.2 (-225; 64.3) |
| **IV** | -3,820 (-12,500; 4,840) | -5,640 (-16,700; 5,380) | -32.4 (-80; 15.2) | -52.1 (-140; 35.3) | 20.8 (-222; 264) | -183 (-399; 32.6) | -382 (-821; 57) | 107 (-25.9; 240) |
| **ST** | -827 (-13,300; 11,600) | 11,600 (-4,040; 27,200) | 1.12 (-67.9; 70.1) | 75.3 (-49.9; 201) | -48 (-395; 299) | 61.4 (-252; 375) | 607 (-16.3; 1,230) | 113 (-78.7; 305) |

**Supplementary Table 2**. Relationship between RAR parameters and cortical thickness

|  | **Frontal lobe** | **Parietal lobe** | **Temporal lobe** | **Occipital lobe** | **Insula** |
| --- | --- | --- | --- | --- | --- |
|  | **B (95% CI)** | **B (95% CI)** | **B (95% CI)** | **B (95% CI)** | **B (95% CI)** |
| **kRA** | 0.611 (-0.185; 1.407) | 0.253 (-0.249; 0.755) | 0.364 (-0.334; 1.062) | 0.107 (-0.128; 0.342) | 0.002 (-0.075; 0.080) |
| **Mesor** | 0.553 (-0.321; 1.427) | 0.192 (-0.359; 0.743) | 0.561 (-0.196; 1.318) | 0.141 (-0.116; 0.398) | 0.048 (-0.035; 0.132) |
| **Amplitude** | 0.542 (-0.346; 1.430) | 0.330 (-0.225; 0.885) | 0.679 (-0.083; 1.441) | 0.216 (-0.041; 0.473) | 0.045 (-0.040; 0.130) |
| **R2** | 0.161 (-0.854; 1.176) | 0.270 (-0.361; 0.901) | 0.426 (-0.450; 1.302) | 0.272 (-0.016; 0.560) | -0.007 (-0.104; 0.090) |
| **M10** | 0.492 (-0.263; 1.247) | 0.249 (-0.223; 0.721) | 0.509 (-0.144; 1.162) | 0.081 (-0.140; 0.303) | 0.069 (-0.002; 0.140) |
| **L5** | 1.000 (0.093; 1.907) | 0.395 (-0.183; 0.973) | 0.801 (0.007; 1.595) | 0.277 (0.010; 0.544) | 0.031 (-0.058; 0.121) |
| **RA** | -0.456 (-1.338; 0.426) | -0.124 (-0.679; 0.431) | -0.271 (-1.041; 0.499) | -0.134 (-0.391; 0.123) | 0.014 (-0.071; 0.099) |
| **IS** | 0.323 (-0.508; 1.154) | 0.255 (-0.262; 0.772) | 0.382 (-0.337; 1.101) | 0.043 (-0.200; 0.286) | 0.056 (-0.023; 0.135) |
| **IV** | -0.611 (-1.371; 0.149) | -0.253 (-0.733; 0.227) | -0.509 (-1.171; 0.153) | -0.027 (-0.253; 0.198) | -0.065 (-0.137; 0.008) |
| **ST** | -0.426 (-1.531; 0.679) | -0.020 (-0.712; 0.672) | 0.196 (-0.766; 1.158) | -0.133 (-0.454; 0.188) | -0.003 (-0.109; 0.103) |

**SUPPLEMENTARY FIGURES**

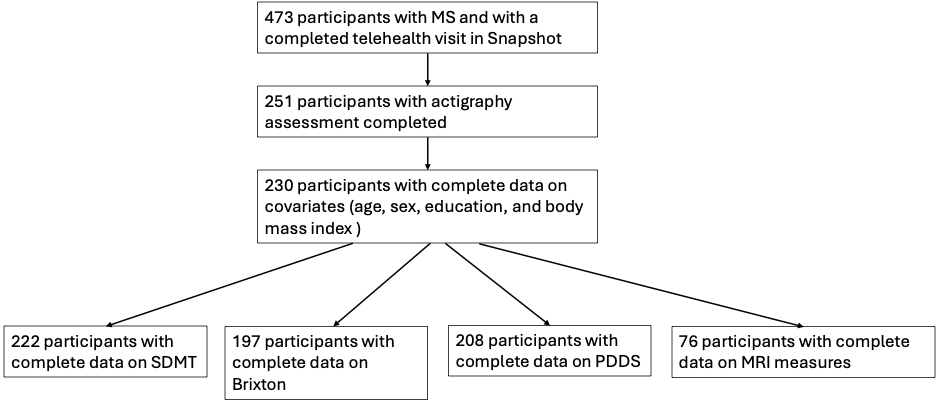

**Supplementary Figure 1**. Sample selection flowchart
